# Variability of enteric pathogen infections by season and meteorological conditions in a low-income, urban setting in Mozambique

**DOI:** 10.1101/2025.10.02.25337035

**Authors:** Rebecca S. Kann, Joshua Garn, Christine S. Fagnant-Sperati, Jedidiah S. Snyder, Erin Kowalsky, Joe Brown, Sandy McGunegill, Rassul Nalá, Matthew C. Freeman, Karen Levy, The PAASIM Study Authorship Consortium

## Abstract

Enteric pathogen transmission is influenced by seasonality and meteorological conditions, yet these relationships are not well understood. We investigated the relationships between (1) season, (2) heavy rainfall events, and (3) temperature and enteric pathogen infections among 12-month-old children in a low-income, urban setting. We analyzed household data and stool samples from 630 participants enrolled in the PAASIM Study in Beira, Mozambique collected between February 2022 and November 2023. We applied generalized estimating equations with robust standard errors to determine the association between season and weather predictors and enteric pathogen prevalence (modified Poisson (PR) for binary outcomes and linear regression (β) for continuous outcomes). During the rainy season, compared to the dry season, we found a 34% lower prevalence of protozoan infections [aPR: 0.66; 95% CI: (0.51, 0.86)], an 11% lower prevalence of co-infections [aPR: 0.89; 95% CI: (0.78, 1.00)], and lower total number of concurrent infections per individual [aβ: –0.17; 95% CI: (–0.38, 0.04)], but no meaningful difference in prevalence of bacterial infections [aPR: 0.97; 95% CI: (0.89, 1.06)] or viral infections [aPR: 0.98; 95% CI: (0.79, 1.22)]. We also saw some associations between rainy season and individual pathogen infections, including a higher prevalence of enteroaggregative *E. coli* (EAEC) [aPR: 1.13; 95% CI: (0.96, 1.33)], enterotoxigenic *E. coli* (ETEC) [aPR: 1.48; 95% CI: (0.88, 2.47], and norovirus infections [aPR: 1.48; 95% CI: (0.93, 2.37)] and a lower prevalence of diffusely adherent *E. coli* (DAEC) [aPR: 0.91; 95% CI: (0.83, 1.00)], atypical enteropathogenic *E. coli* (aEPEC) [aPR: 0.88; 95% CI: (0.74, 1.05)], and *Cryptosporidium* [aPR: 0.40; 95% CI: (0.27, 0.60)] during the rainy season. Following heavy rainfall events (1-week lag), there was a 30% higher prevalence of protozoan infections [aPR: 1.30; 95% CI: (1.06, 1.59)], a 22% higher prevalence of viral infections [aPR: 1.22; 95% CI: (0.95, 1.57)], and a 10% higher prevalence of co-infections [aPR: 1.09; 95% CI: (0.99, 1.21)], but no meaningful difference in prevalence for bacterial infections [aPR: 1.01; 95% CI: (0.92, 1.10)] and no difference in the total number of infections [aβ: –0.04; 95% CI: (–0.22, 0.14)]. Temperatures above the median (1-week lag), compared to below the median, were associated with a 35% lower prevalence of protozoan infections [aPR: 0.65; 95% CI: (0.49, 0.86)] and a 14% lower prevalence of co-infections [aPR: 0.86; 95% CI: (0.76, 0.97)], but we did not find evidence of a difference in bacterial infections [aPR: 0.98; 95% CI: (0.89, 1.07)], viral infections [aPR: 0.94; 95% CI: (0.74, 1.19)], or the total number of infections [aβ: –0.13; 95% CI: (–0.33, 0.07)] by temperature. Our results contrast many previous studies that have predominately shown a higher risk of bacterial infections and a lower risk of viral infection during periods with higher temperatures and precipitation but align with previous research suggesting a higher prevalence of some enteric infections following heavy rainfall events. Both long-term seasonal trends in enteric infections as well as the more immediate effects of extreme weather events, such as heavy rainfall events, are therefore important for designing interventions.

## Introduction

Diarrheal disease, often caused by enteric pathogen infections, is a leading cause of under-5 child morbidity and mortality in low– and middle-income countries.^1^ Diarrheal disease and enteric pathogen transmission pathways have distinct seasonal patterns and are driven by different meteorological conditions, including temperature and precipitation.^2–4,4,5^ These seasonal variations can be attributed to changes in environmental conditions and human behaviors.^3,6^ Because of the well-documented influences of seasonal and meteorological conditions on enteric pathogen transmission patterns, interventions aimed at reducing the burden of enteric diseases need to consider the role of these factors to develop optimal intervention strategies.

The environmental transmission of enteric pathogens varies by season and is influenced by specific weather conditions such as temperature and precipitation.^3,7^ Temperature impacts pathogen survival and reproduction in the environment, with some pathogens, such as bacteria and protozoa, showing higher transmission and virulence at higher temperatures and other pathogens, including viruses, having peaks at lower temperatures.^3^ Precipitation influences pathogen mobility and distribution, with light to moderate rainfall leading to the distribution of pathogens in the environment, and high levels of rainfall leading to a flushing effect where pathogens are washed into water sources.^3,8,9^ Droughts are also important as they can lead to a higher risk of exposures due to the accumulation and concentration of pathogens in the environment and a higher exposure risk when heavy rainfall events (HREs) subsequently wash out concentrated contaminants.^8–10^ The effects of these various meteorological conditions are complex and impact enteric pathogen transmission pathways through both direct changes in environmental conditions as well as indirect changes in human behaviors and local contexts.

When studying the effects of temperature and precipitation on enteric health, it is important to consider the differences in the survival and transmissibility of specific pathogens through the environment. In general, viral infections often peak in cooler conditions, whereas bacterial and protozoan pathogens exhibit greater persistence and transmission in warmer conditions, particularly in water and animal reservoirs.^3,11,12^ However, many previous studies of the relationship between climate and health have been limited by the use of reported diarrhea as the primary indicator of enteric health, which is prone to bias, cannot detect asymptomatic infections, and is non-cause specific.^7,13,14^ More recent studies using data on enteric pathogen infections have shown distinct pathogen-specific effects of seasonal patterns and weather extremes on enteric infections.^2,5,15,16^ Data on specific enteric pathogen infections, rather than just reported diarrhea, may be important for illuminating taxa– and pathogen-specific differences in seasonal variability in transmission pathways, which is important for developing targeted interventions.

Seasonal variation in enteric pathogen transmission is also influenced by socioeconomic conditions and human behaviors.^3,17–20^ Previous studies have shown variation in the impacts of weather on enteric pathogens across different countries, climates, and demographic contexts.^2,10^ For example, during long periods of drought, people may be more likely to use unsafe alternate water sources or store their water for longer periods of time, leading to a higher risk of enteric pathogen exposures.^20^ During outbreaks, disease prevention behaviors, including behaviors widely adopted during the COVID-19 pandemic—handwashing and social distancing—have the potential to disrupt transmission patterns, though water, sanitation, and hygiene insecurity may reduce their effectiveness.^21–24^ It is important to understand these context-specific variations in weather-driven changes in human behaviors in order to develop targeted interventions.

The PAASIM study (“*Pesquisa sobre o Acesso à Água e a Saúde Infantil em Moçambique” –* Research on Access to Water and Child Health in Mozambique) may provide valuable context for understanding the ways weather and seasonality impact enteric pathogen transmission. Most previous studies of the effects of weather on enteric disease have been focused in rural^15,25–28^ or high-income settings,^29–37^ or they have relied on reported diarrhea or other reported measures of gastrointestinal illness.^10,38–42^ Multi-country studies have shown distinct differences in enteric infections across different contexts,^2,16,38^ including one study across different contexts in Mozambique, which showed that coastal regions had distinct seasonal patterns of diarrheal disease that were different than other areas of Mozambique.^41^ Previous analyses from the PAASIM study have shown that weather and seasonality impact water quality and intermittency, with some evidence suggesting an association between seasonality and diarrhea [Hubbard et al., and Kann et al., *manuscripts in preparation*]. However, prior PAASIM analyses revealed no association between specific meteorological conditions, including HREs and ambient temperature, and diarrhea, potentially due to the limitations of reported diarrhea as an outcome measure.^43^ Data on enteric pathogen infections from the PAASIM study can add insight into seasonal and weather-driven patterns in enteric disease that have the potential to be generalizable to other coastal, urban contexts.

In this study, we assessed how season and meteorological conditions impacted the prevalence of taxa-specific enteric pathogen infections for children in Beira, Mozambique. Specifically, we examined relationships between (1) seasonality, (2) heavy rainfall events, and (3) temperature on enteric pathogen infections among 12-month-old children enrolled in the PAASIM study cohort. Our findings provide insight into how seasonal changes and weather extremes may influence enteric infection patterns in similar low-income, urban settings, which will be valuable for developing targeted interventions aimed at reducing the burden of enteric disease in similar contexts.

## Methods

### Study setting

Mozambique is located on the southeastern coast of Africa and much of its population is concentrated on its expansive coastline in urban areas.^44,45^ Mozambique is one of the most climate-vulnerable countries in the world^46^ and Beira City is particularly vulnerable due to rising sea levels and coastal erosion which puts large parts of the city at risk of regular flood events and infrastructure vulnerability.^46–48^ Diarrhea is a leading cause of mortality in Mozambique, with children under 5 years old facing the highest burden.^49^

### Study design and data collection

We used data from the PAASIM study, a cluster-matched study evaluating the impact of a large-scale urban piped water supply improvement on children’s enteric pathogen infections and other health outcomes in Beira, Mozambique.^50^ Mothers were recruited and enrolled in the study in their third trimester of pregnancy between February 19^th^, 2021and October 7^th^, 2022. Mother-child dyads were followed through four follow-up visits, when the child was 3-, 6-, 9-, and 12-month-old. Details on the participant recruitment, eligibility, and enrollment for the PAASIM study can be found elsewhere.^51^ This analysis was conducted as an observational study, independent of the water supply intervention.

We used data collected during the 12-month-old visits for the PAASIM study, which occurred between February 2022 and November 2023. In total, 642 mother-child dyads completed all visits from pre-birth through 12-months. We excluded households that did not have any enteric pathogen infection data available at the 12-month visit (n=12), for a final sample size of 630 households. During the 12-month visits, enumerators collected household surveys and a stool sample from the child. Study staff conducted household surveys in Portuguese and entered responses electronically into password protected tablets via the Open Data Kit (ODK) platform.^52^

### Exposure: Season and weather variables

Season was dichotomized into rainy vs. dry season, where surveys collected between October and April correspond to the rainy season and surveys collected between May and September correspond to the dry season. We used a data driven approach, described by Chao et al.,^2^ which included conducting a principal component analysis (PCA) and k-means clustering to group months into different seasons based on their similarity by weather conditions. We included daily data on rainfall, temperature, specific humidity and relative humidity in the PCA. We then implemented k-means clustering using the first 3 components from the PCA to group months by similarity. Our data driven seasons aligned with general trends in temperature and rainfall, where the rainy season (October-April) is typical of higher rainfall and higher temperatures and the dry season (May-September) is typical of lower rainfall and lower temperatures (Figure 1C).

**Figure 1.**
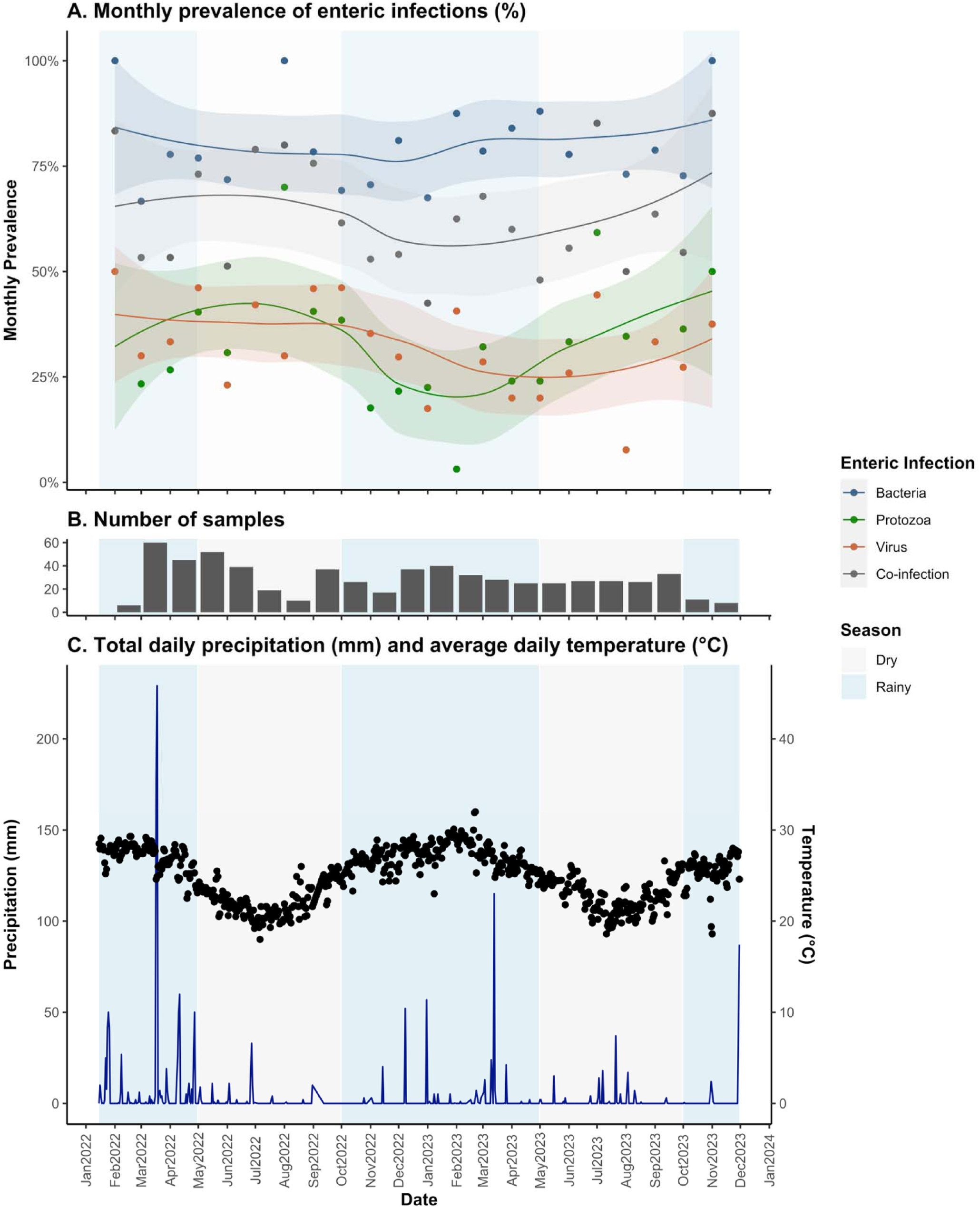
Summary of enteric infections and weather throughout study period for the PAASIM study. For all plots, rainy season is shaded in blue and dry season is shaded in grey. (A) Monthly prevalence of enteric infections by taxa with Loess curves and 95% confidence interval in shaded area. (B) Total number of samples collected per month, where the earliest sample collection date was February 17^th^, 2022, and the last collection date was November 3, 2023. (C) Sum of total daily precipitation (blue line) and daily average temperature (black points). All weather data was collected from the NOAA GSOD land surface station at the Beira ational Airport.

**Figure 2.**
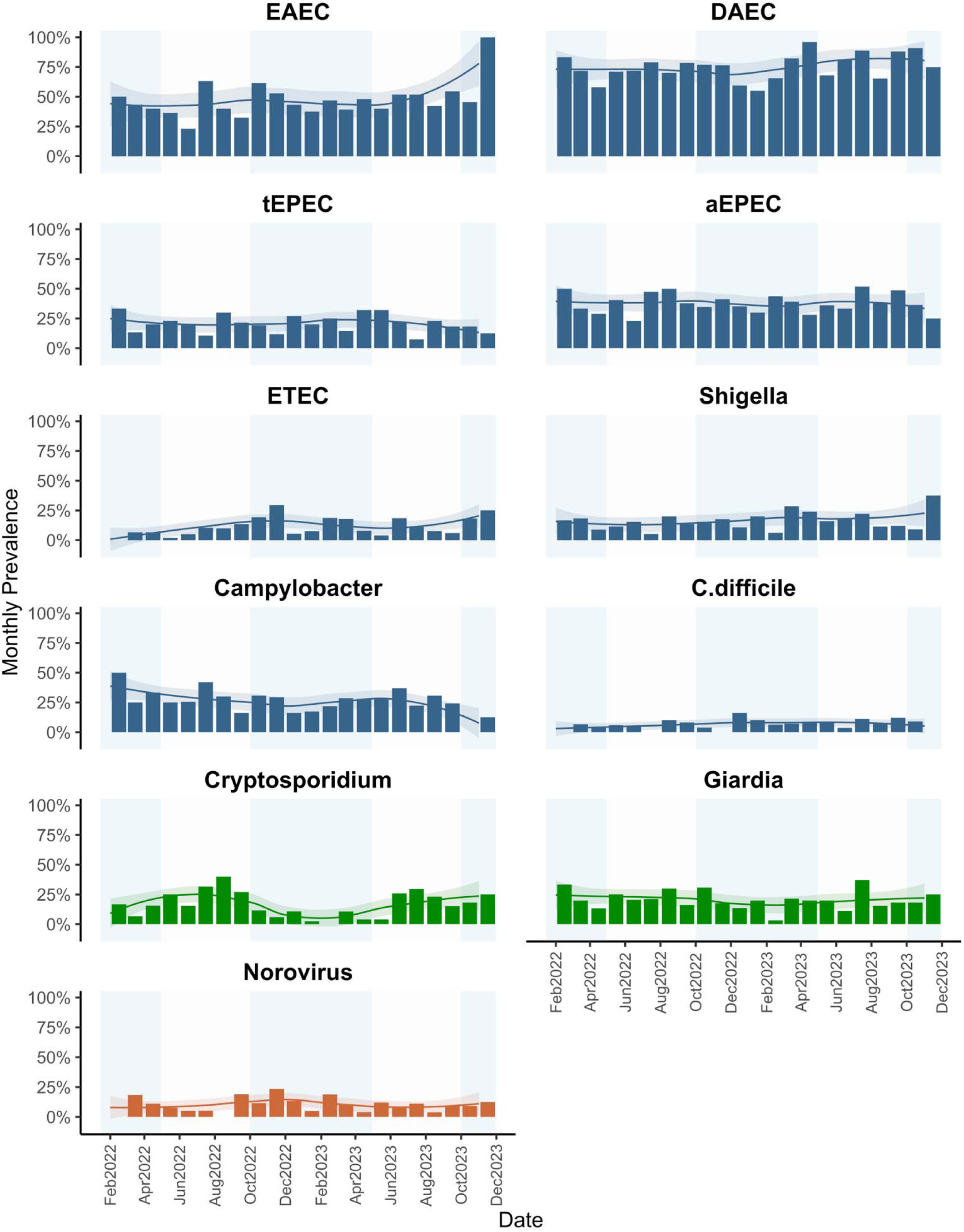
Monthly prevalence of individual pathogen infections with Loess curves and 95% confidence interval in shaded area. Individual enteric infection outcomes only summarized for those with overall prevalence>10%.

Heavy rainfall events (HRE) were defined as total daily rainfall above the 95^th^ percentile (8.81mm) for the overall study period. HREs were considered weather events of interest for higher risk of exposure due to the higher likelihood of infrastructure damage and flooding following HREs, as has been shown in previous literature.^10,15,30,35^ We also ran a sensitivity analysis assessing HRE cutoff points of 90^th^ and 80^th^ percentiles, for comparability with prior studies.^10,15,30,35^ We assessed the effects of temperature using a categorical variable to compare the effects of rolling average weekly temperatures above vs. below the median temperature (25.0°C) for the full study period. We also ran a sensitivity analysis assessing the effects of rolling average weekly temperatures above vs. below the bottom tertile (23.3°C) and the upper tertile (26.3°C) for the full study period. We assessed the effects of HREs and above median temperatures at 3 lag periods – 0 week (corresponding to the period 0-1 weeks before sample collection), 1 week (1-2 weeks before sample collection), and 2 weeks (2-3 weeks before sample collection), based on previous studies which have shown these to be relevant periods of exposure between weather events and enteric health outcomes.^15,16,53^

We assessed antecedent conditions, or the conditions preceding an HRE, as a modifier of the effects of HREs on enteric infections based on previous studies that have shown a higher risk of HREs following dry periods.^8–10^ Using methods described by Carlton et al., 2014, antecedent weather conditions were calculated using the sum of total rainfall over the 8 weeks prior to sample collection.^8^ Wet conditions are those exceeding the 67^th^ percentile, medium conditions were conditions within the 67^th^ percentile and the 33^rd^ percentile, and dry conditions were those below the 33^rd^ percentile for total rainfall over the 8-week period. There were no instances of HRE events following dry conditions, so we dichotomized the antecedent conditions in models to compare wet conditions to medium or dry conditions.

Weather data was obtained from the National Oceanic and Atmospheric Administrations (NOAA) Global Surface Summary of the Day (GSOD) database, which included data from a land surface station at the Beira International Airport,^54^ located approximately seven kilometers from the study area, using the “gsodr” R package.^55^ Daily temperature and rainfall measurements were obtained for each day that a survey was collected. Missing weather data (n=11 missing days in the study period) was imputed using a linear trend between the two nearest adjacent dates. Daily precipitation and temperature data were matched to the date of stool sample collection.

### Outcome: Enteric Pathogen Infections

We used both combined and individual measures of enteric pathogen infection data to assess health outcomes in this study. Combined outcome measures included the prevalence of any bacterial pathogen, any protozoan pathogen, any viral pathogen, co-infection (more than one pathogen), and the total number of pathogens detected in stool. Enteroaggregative *E. coli* (EAEC) and diffuse adherent *E. coli* (DAEC) were excluded from combined outcome measures because there is mixed evidence about their pathogenicity and clinical relevance.^56–61^ We assessed relationships with individual pathogen prevalence for pathogens with a prevalence over 10%, which included EAEC, DAEC, typical enteropathogenic *E. coli* (tEPEC), atypical enteropathogenic *E. coli* (aEPEC), enterotoxigenic *E. coli* (ETEC), *Shigella spp./*enteroinvasive *E. coli* (EIEC), *Campylobacter jejuni*/*C. coli*, norovirus, *Cryptosporidium spp.*, and *Giardia duodenalis*. For this study, detection of pathogens in stool (presence/absence) is used as an indicator of infection.^62^

Stool samples collected from children at the 12-month visit were assayed for enteric pathogen infections using TaqMan Array Card (TAC, ThermoFisher Scientific, Waltham, MA, USA) assays, which use real-time quantitative Polymerase Chain Reaction (qPCR) via a 384-well microfluidic card for simultaneous detection of multiple viral, bacterial, and parasitic enteric pathogen targets as well as antimicrobial resistance genes,^63^ customized for our targets of interest (see Levy et al., 2023 Supplemental Table 2 for full list of pathogens tested by TAC).^51^

### Other covariates

Data for all other covariates included in models were collected during household surveys and were chosen based on *a priori* hypotheses about confounders and precision variables for relationships of interest. Household surveys contained questions and structured observations related to household demographics, infant health, water, sanitation, and hygiene practices, and household conditions. All models adjusted for intervention status (intervention vs. comparison), access to a direct household water connection (presence vs. absence), caregiver’s education level (at least secondary education vs. below secondary education), caregiver’s employment status (fixed employment vs. non-fixed employment), and access to basic sanitation (presence vs. absence). We also adjusted for poverty level in all models, using the Mozambique Simple Poverty Scorecard,^64^ dichotomized to compare high poverty vs. not high poverty households, where households with a score less than 66 on the scorecard were considered high poverty.

### Statistical Analyses

We ran regression models to investigate relationships between (1) seasonality, (2) HREs, and (3) temperature, as exposure variables, on enteric pathogen infection outcomes. We also ran models for the effect of HREs including the interaction between HRE and antecedent conditions, giving estimates of the effect of HREs on enteric infections following (a) wet conditions and (b) dry or medium conditions.

We calculated effect estimates using generalized estimating equations with robust standard errors and an exchangeable correlation matrix to account for clustering at the sub-neighborhood level to obtain prevalence ratios (modified Poisson regression) for binary outcomes^51,52^ and mean differences (linear regression) for continuous outcomes. In addition to confounders that we pre-specified to use in all models (see above), in models assessing the effects of HREs, we adjusted for the rolling 7-day average temperature over the same period. Similarly, for models assessing the effects of above median temperatures, we adjusted for the rolling 7-day average precipitation over the same period. All analyses were conducted using R statistical software (RStudio v. 4.3.1).^65^

### Ethics statement

The PAASIM study was approved by the Mozambique National Bio-Ethics Committee for Health (Ref: 105/CNBS/20) and Emory University’s Institutional Review Board (IRB#: CR001-IRB00098584, Atlanta, GA). Credential letters and permissions were obtained from Beira municipality and municipal district administrations from study neighborhoods for study conduct in those areas. Study participants were given consent forms at their homes at a pre-birth/enrollment visit, which were signed or marked with a thumbprint by the primary caregiver or a parent or guardian over 18 years old.

## Results

### Study population

Demographic and enteric infection data for participants in the PAASIM study are published elsewhere^66^; data for the population sample for this analysis (N=630) are shown in Supplemental Table 1. There is a high level of poverty within this study population, with 383 participants (60.8%) considered high poverty and 297 (47.1%) considered severely food insecure (Supplemental Table 1). Only 129 participants (20.5%) had access to a handwashing station in their household and 226 (35.9%) had basic sanitation access. Despite every participant having access to an improved drinking water source, only 305 (48.4%) had access to a water source on premises, and there was a high level of intermittency in the system, with water only being available for 13.4 hours per day on average.

### Meteorological Conditions

During the period when stool samples were collected, 463 days (73.5%) had 0 mm of rainfall (median = 0 mm, 3^rd^ quartile = 0.25 mm) (Supplemental Figure 1). The maximum total rainfall in one day was 229mm, which was recorded on March 3^rd^, 2022. Temperatures ranged from 18-32°C, with a median temperature of 25.0°C.

#### 1. Season on enteric infections

Rainy season, compared to the dry season, was associated with a 34% lower prevalence of protozoan infections [aPR: 0.66; 95% CI: (0.51,0.86)] and an 11% lower prevalence of co-infections [aPR: 0.89; 95% CI: (0.78, 1.00)] (Table 1). The total number of concurrent infections within individuals was also lower in the rainy season compared to the dry season [aβ: –0.17; 95% CI: (–0.38, 0.04)]. There was no meaningful difference in the prevalence of bacterial infections or viral infections in the rainy season compared to the dry season.

**Table 1.** Prevalence of enteric pathogen infections stratified by season with adjusted PR and β estimates from generalized estimating equation models assessing associations between season (rainy vs. dry) and enteric infections (N=630)

| | Rainy Season<br>[N=335] | Dry Season<br>[N=295] | Adjusted PR or $\beta$<br>(95% CI)* | p-value |
| --- | --- | --- | --- | --- |
| **Combined outcomes** |  |  |  |  |
| Presence of any bacterial infection | 255 (76.1) | 233 (79.0) | 0.97 (0.89, 1.06) | 0.53 |
| Presence of any protozoan infection | 83 (24.8) | 114 (38.6) | 0.66 (0.51, 0.86) | 0.00 |
| Presence of any viral infection | 104 (31.0) | 98 (33.2) | 0.98 (0.79, 1.22) | 0.86 |
| Presence of any co-infection (>1 infection) | 190 (56.7) | 193 (65.4) | 0.89 (0.78, 1.00) | 0.06 |
| Number of individual pathogens detected | 1.49 (1.13) | 1.69 (1.12) | -0.17 (-0.38, 0.04) | 0.10 |
| **Individual bacterial infections** |  |  |  |  |
| Enteroaggregative *E. coli* (EAEC) | 154 (46.0) | 123 (41.7) | 1.13 (0.96, 1.33) | 0.14 |
| Diffusely adherent *E. coli* (DAEC) | 235 (70.1) | 225 (76.3) | 0.91 (0.83, 1.00) | 0.06 |
| Shiga toxin-producing *E. coli* (STEC) infection | 335 (100.0) | 295 (100.0) | -- | -- |
| Enteropathogenic *E. coli* (tEPEC) infection | 67 (20.0) | 61 (20.7) | 0.99 (0.72, 1.36) | 0.95 |
| Enteropathogenic *E. coli* (aEPEC) infection | 115 (34.3) | 116 (39.3) | 0.88 (0.74, 1.05) | 0.17 |
| Enterotoxigenic *E. coli* (ETEC) infection | 39 (11.6) | 24 (8.1) | 1.48 (0.88, 2.47) | 0.14 |
| Shigella spp./Enteroinvasive *E. coli* (EIEC) infection | 55 (16.4) | 42 (14.2) | 1.21 (0.79, 1.87) | 0.39 |
| *E. coli* O157 infection | 10 (3.0) | 10 (3.4) | -- | -- |
| Campylobacter jejuni/C. coli infection | 82 (24.5) | 79 (26.8) | 0.96 (0.72, 1.27) | 0.76 |
| Salmonella spp. infection | 6 (1.8) | 6 (2.0) | -- | -- |
| Vibrio cholerae infection | 8 (2.4) | 5 (1.7) | -- | -- |
| Clostridium difficile infection | 24 (7.2) | 21 (7.1) | -- | -- |
| **Individual viral infections** |  |  |  |  |
| Adenovirus infection | 20 (6.0) | 7 (2.4) | -- | -- |
| Astrovirus infection | 14 (4.2) | 19 (6.4) | -- | -- |
| Norovirus infection | 42 (12.5) | 26 (8.8) | 1.48 (0.93, 2.37) | 0.10 |
| Rotavirus infection | 13 (3.9) | 32 (10.8) | -- | -- |
| Sapovirus infection | 30 (9.0) | 29 (9.8) | -- | -- |
| SARS-CoV-2 infection | 3 (0.9) | 4 (1.4) | -- | -- |
| **Individual protozoan infections** |  |  |  |  |
| Cryptosporidium infection | 29 (8.7) | 66 (22.4) | 0.40 (0.27, 0.60) | 0.00 |
| Giardia infection | 60 (17.9) | 62 (21.0) | 0.88 (0.63, 1.21) | 0.44 |
| Entamoeba histolytica infection | 335 (100.0) | 295 (100.0) | -- | -- |
| Cyclospora cayetanensis infection | 0 (0.0) | 1 (0.3) | -- | -- |

\*All models adjusted for intervention status, access to a direct household connection to a piped water source, poverty, caregiver education level, caregiver employment status, and basic sanitation access. Models for individual enteric infection outcomes only assessed for infections with prevalence > 10%.

There was a higher prevalence in the rainy season compared to the dry season of EAEC [aPR: 1.13; 95% CI: (0.96, 1.33)], ETEC [aPR: 1.48; 95% CI: (0.88, 2.47)], and norovirus [aPR: 1.48; 95% CI: (0.93, 2.37)] infections, but a lower prevalence of DAEC [aPR: 0.91; 95% CI: (0.83, 1.00)], aEPEC [aPR: 0.88; 95% CI: (0.74, 1.05)], and *Cryptosporidium* [aPR: 0.40; 95% CI: (0.27, 0.60)] infections. There was no meaningful difference in the prevalence of tEPEC, *Shigella*, *Campylobacter*, or *Giardia* when comparing rainy season to dry season.

#### 2. HREs on enteric infections

When comparing HREs 1-2 weeks before sample collection to no HREs, there was a higher prevalence of protozoan infections [aPR: 1.30; 95% CI: (1.06, 1.59)], viral infections [aPR: 1.22; 95% CI: (0.95, 1.57)], and co-infections [aPR: 1.09; 95% CI: (0.99, 1.21)] (Figure 3, Supplemental Table 2). There was also a higher prevalence of viral infections following HREs 0-1 week before sample collection, compared to no HREs [aPR: 1.24; 95% CI: (0.97, 1.60)]. There was no meaningful difference in the prevalence of infections when comparing HREs to no HREs for bacterial infections at any lag period, for protozoan infections at the 0-week and 2-week lag periods, for viral infections at the 2-week lag period, for co-infections at the 0-week or 2-week lag periods, or for the total number of infections at any lag period.

**Figure 3.**
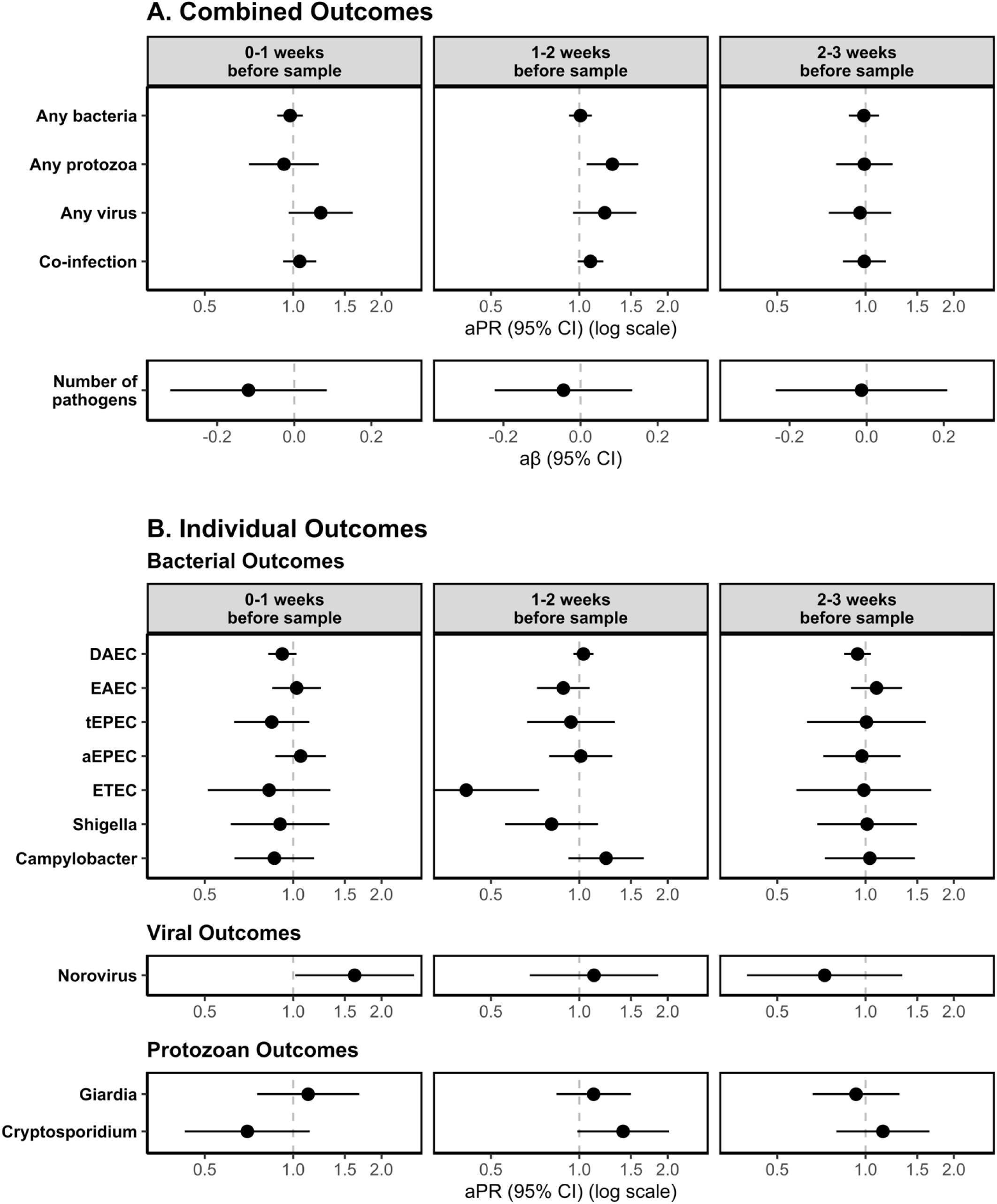
Adjusted associations of Heavy Rainfall Events (HREs) on enteric pathogen infections by enteric pathogen outcome type (row) and lag period (column) for. (A) combined outcome measures and (B) individual outcome measures (with prevalence >10%). HREs were defined as a day where the total rainfall was above the 95th percentile (8.81mm) for the overall study period. All models adjusted for rolling mean temperature during the same period, intervention status, access to a direct household connection to a piped water source, poverty, caregiver education level, caregiver employment status, and basic sanitation access.

For models assessing the effects of HREs on individual enteric pathogen infections, we found a higher prevalence of norovirus infections [aPR: 1.62; 95% CI: (1.02, 2.58)], and a lower prevalence of DAEC infections [aPR: 0.92; 95% CI: (0.82, 1.03)] following HREs 0-1 week before sample collection, compared to no HREs in that period. We also found a higher prevalence of *Cryptosporidium* infections [aPR: 1.41; 95% CI: (0.98, 2.02)] and a lower prevalence of ETEC infections [aPR: 0.41; 95% CI: (0.23, 0.73)] following HREs 1-2 weeks before sample collection, compared to no HREs in that period. We did not find evidence of a difference in the risk of DAEC, ETEC, norovirus, or *Cryptosporidium* following HREs at other lag periods. We also did not see a meaningful difference in the prevalence of EAEC, tEPEC, aEPEC, *Shigella*, *Campylobacter*, or *Giardia* following HREs at any lag period. Models for the effect of HREs at different cutoff points (80^th^ percentile and 90^th^ percentile) show similar trends though associations were slightly attenuated towards the null at lower percentiles (Supplemental Table 3).

Our analysis of the effect of antecedent conditions as a modifier of the effects of HREs on enteric infections showed evidence of only two models with meaningful interaction between wet and medium/dry antecedent conditions (Figure 4, Supplemental Table 4). For HREs 2-3 weeks before sample collection, compared to no HREs, the prevalence of viral infections was higher following wet conditions, compared to dry conditions [Interaction term p-value: 0.01] and the prevalence of DAEC infections was also higher following wet conditions, compared to dry conditions [Interaction term p-value: 0.01]. (Figure 4, Supplemental Table 4).

**Figure 4.**
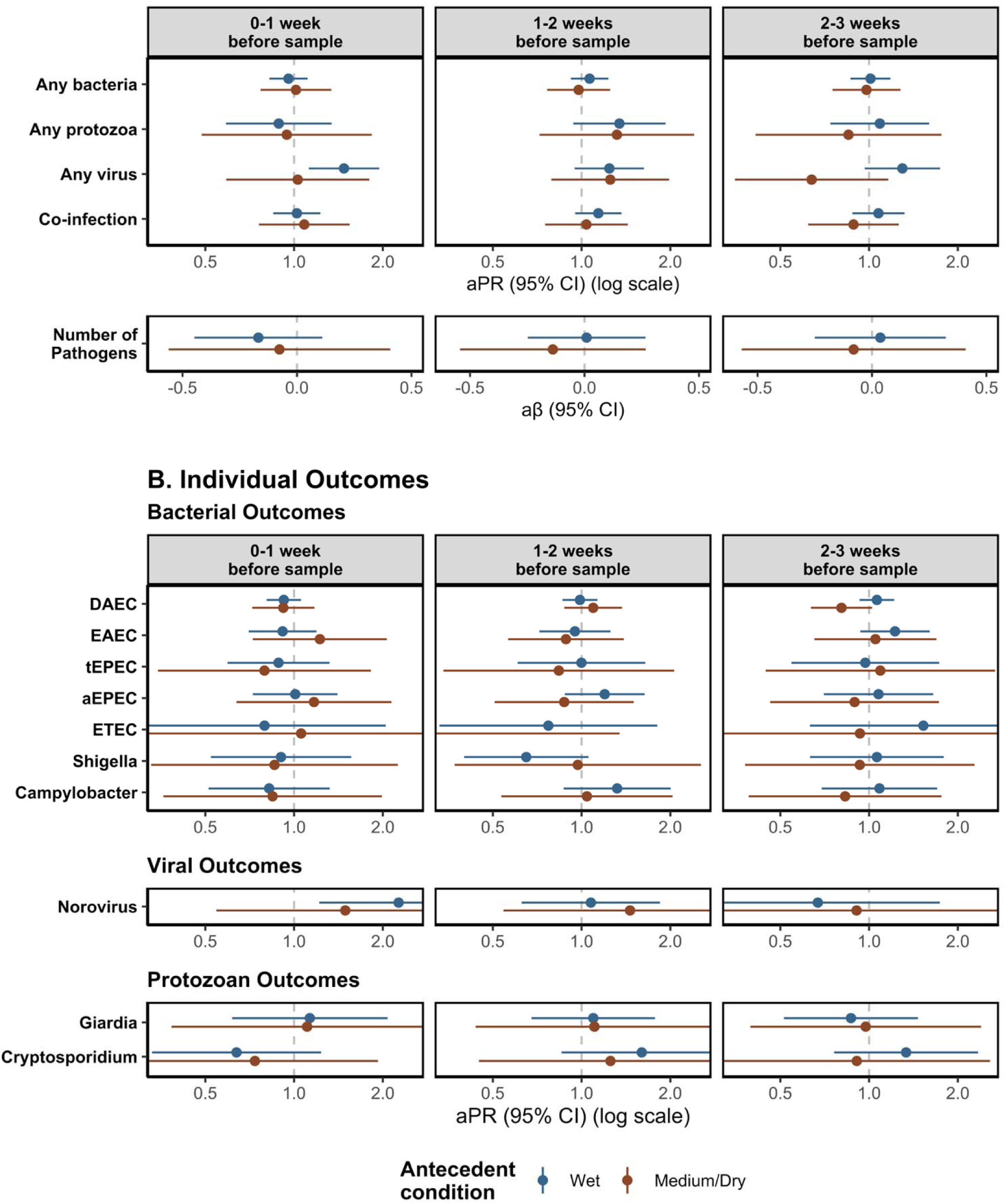
Adjusted associations of Heavy Rainfall Events (HREs) on enteric pathogen infections including interaction with antecedent conditions, by enteric pathogen outcome type (rows) and lag period (columns). Results stratified by antecedent conditions. Antecedent condition was calculated using the sum of total rainfall over the 8 weeks prior to e collection, where wet conditions are those exceeding 67^th^ percentile for total rainfall over the 8-week period compared to the study period and medium/dry conditions are those below the 67^th^ percentile for total rainfall. HREs were defined as a day where the total rainfall was above the 95^th^ percentile (8.81mm) for the overall study period. All models adjusted for rolling mean temperature during the same period, intervention status, access to a direct household connection to a piped water source, poverty, caregiver education level, caregiver employment status, and basic sanitation access. Numeric results shown in Supplemental Table 4.

#### 3. Temperature on enteric infections

When comparing above median temperatures to below median temperatures, there was a lower prevalence at all lag periods of protozoan infections [0-week lag: aPR: 0.62; 95% CI: (0.47, 0.80); 1-week lag: aPR: 0.65; 95% CI: (0.49, 0.86); 2-week lag: aPR: 0.62; 95% CI: (0.48, 0.79)] and co-infections [0-week lag: aPR: 0.91; 95% CI: (0.80, 1.04); 1-week lag: aPR: 0.86; 95% CI: (0.76, 0.97); 2-week lag: aPR: 0.92; 95% CI: (0.81, 1.03)] (Figure 5, Supplemental Table 5). There was no meaningful difference in the prevalence of bacterial infections or viral infections, or the total number of infections when comparing above to below median temperatures at any lag period.

**Figure 5.**
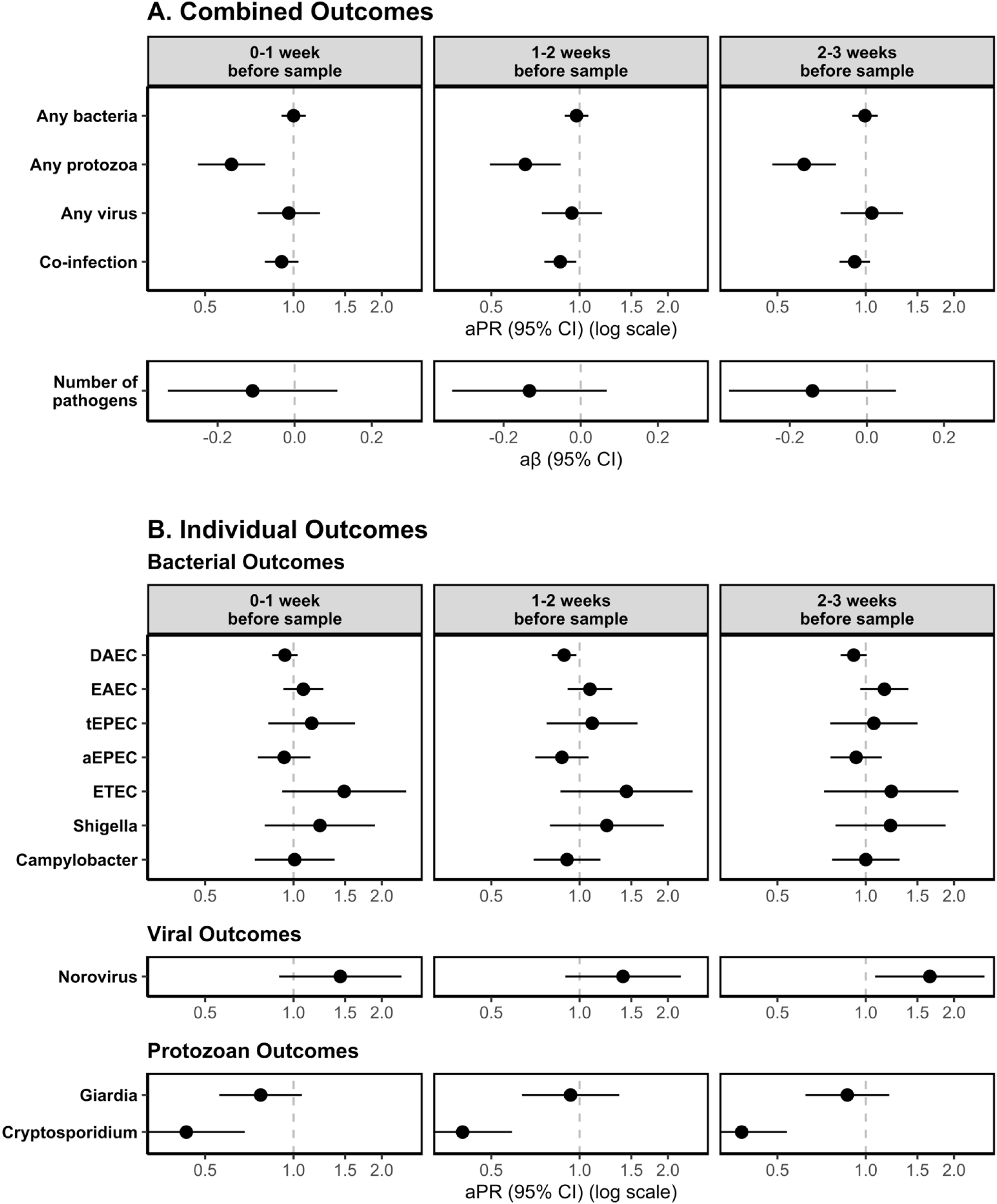
Adjusted associations of above median temperatures on enteric pathogen infections by enteric pathogen outcome type (row) and lag period (column) for (A) combined outcomes and (B) individual enteric pathogen outcomes. Above median temperatures were defined as rolling average weekly temperature above the 50th percentile (25.0°C) for the full study period. All models adjusted for rolling mean precipitation during the same period, intervention status, access to a direct household connection to a piped water source, poverty, caregiver education level, caregiver employment status, and basic sanitation access.

When assessing the effects of temperature on individual enteric pathogen infections, we found a lower prevalence of DAEC infections following above vs. below median temperatures 1-2 weeks before sample collection [aPR: 0.89; 95% CI: (0.80, 0.98)] and 2-3 weeks before sample collection [aPR: 0.91; 95% CI: (0.82, 1.01)]. At all lag periods, above vs. below median temperatures were associated with a higher prevalence of norovirus infections [0-week lag: aPR: 1.43; 95% CI: (0.89, 2.33); 1-week lag: aPR: 1.41; 95% CI: (0.89, 2.21); 2-week lag: aPR: 1.65; 95% CI: (1.08, 2.54)] and a lower prevalence of *Cryptosporidium* infections [0-week lag: aPR: 0.43; 95% CI: (0.27, 0.68); 1-week lag: aPR: 1.41; 95% CI: (0.89, 2.21); 2-week lag: aPR: 0.38; 95% CI: (0.27, 0.54)]. There was also a higher prevalence of ETEC infections [aPR: 1.49; 95% CI: (0.92, 2.42)] and a lower prevalence of *Giardia* infections [aPR: 0.77; 95% CI: (0.56, 1.07)] following above median temperatures 0-1 week before sample collection. We did not find evidence of a difference in prevalence when comparing above vs. below median temperatures for other individual enteric pathogen infections. Models for the effect of average weekly temperatures below the lower tertile and above the upper tertile show similar trends (Supplemental Tables 6 & 7).

## Discussion

Our results show seasonal and weather-dependent trends in enteric pathogen infections prevalence in a low-income, coastal, urban setting. We found a lower prevalence of protozoan infections, co-infections, and total number of infections, during the rainy season. We also found associations between seasonality and some individual pathogens. Specifically, there was a higher prevalence of EAEC, ETEC, and norovirus infections during the rainy season whereas there was a higher prevalence of DAEC, aEPEC, and *Cryptosporidium* infections during the dry season. HREs (1-week lag period) were associated with a higher prevalence of protozoan infections, viral infections, and co-infections as well as a higher prevalence of norovirus and *Cryptosporidium* infections and a lower prevalence of ETEC and DAEC infections. Higher temperatures were associated with a lower prevalence of protozoan infections and co-infections as well as a lower prevalence of DAEC and Cryptosporidium infections and a higher prevalence of ETEC and norovirus infections.

Our findings of a harmful association of HREs on protozoan infections, viral infections, and co-infections, aligns with relationships shown in previous literature. HREs can flush environmental contaminants into waterways, which can lead to contamination of drinking water and a higher risk of enteric pathogen exposures.^3,67^ HREs have been shown to be more harmful following periods of drought.^8,10^ We did not find consistent differences in the effect of HREs by antecedent conditions, potentially because there were not any HREs following dry conditions, so we were only able to compare wet conditions to dry or medium conditions. Compared to seasonality, which is dependent on long term trends in meteorological conditions, HREs represent a more proximal exposure with the potential to have more immediate impacts on enteric infections. Whereas our results assessing the effects of seasonality were impacted by more long-term trends in the specific context of the study period, the effects of HREs more closely aligned with previous research, regardless of the context, likely due to HREs acting as a more proximal level of exposure. Our results align with conclusions made in previous research suggesting a need for interventions to prioritize periods following HREs.

Interestingly, our results showed seasonal patterns of enteric pathogen infections that differed from most previous literature in this field. While most previous research has shown higher risks of bacterial and protozoan infections during rainy season and at higher temperatures,^3,68^ we found a lower risk of protozoan infections, co-infections, and the total number of infections during the rainy season and no association between season and viral infections or bacterial infections. One of the reasons we may not see variation in prevalence of bacterial infections is that there was a very high overall prevalence of infections in the study area (>77%), while many previous studies have shown comparatively lower prevalence of infection and diarrhea.^10,15,26,27^ Given such high force of infection, it may not have been possible for the prevalence to further increase under high-risk conditions (i.e., rainy season), and transmission may have been continually sustained in the lower-risk conditions (i.e., dry season). On the other hand, we did see evidence of seasonal trends in protozoan infections, co-infections, and the total number of infections, which were potentially influenced by the context of data collection following two humanitarian emergencies – a large cyclone in the area and the acute phase of the COVID pandemic.^69^ A previous analysis in the PAASIM study area showed that there was a high level of adherence to disease prevention behaviors, including handwashing and social distancing, and that these behaviors were dependent on people’s WASH access.^21^ It is possible there was variation in people’s ability to practice disease prevention behaviors based on seasonal variation in water availability. For example, handwashing may have been less accessible, leading to a higher risk of exposure, during the dry season when water intermittency was higher in the study area [Hubbard et al., *manuscript in preparation*]. Additionally, damage to infrastructure as well as an influx in investment in infrastructure in the area caused by Cyclone Idai likely impacted people’s WASH access and influenced infection trends in the study area in a way that was different than previous studies. Finally, our results may be limited by the assessment of the prevalence, not incidence, of enteric infections in stool as the primary health outcome. With these data, we are unable to distinguish between new and long-term infections in the study population.^62^ In areas with high force of infection, the prevalence of enteric infections may not vary by season, and disease outbreaks, or other humanitarian emergencies, may influence unexpected long-term disease patterns.

We observed a lower prevalence of protozoan infections and co-infections at higher temperatures at all lag periods and no association between temperature and viral or bacterial infections or the total number of infections, contradicting with previous research. Evidence from previous studies has shown that higher temperatures are associated with bacterial infections, whereas lower temperatures are associated with viral infections.^17,38^ Previous assessments of the effect of temperature on protozoan infections are mixed, with some showing a higher risk of *Cryptosporidium* and *Giardia* with higher temperatures^16^ and others showing no effect of temperature.^15,70^ In this study area, temperature and seasonality were highly correlated, suggesting associations with temperature may be at least partially driven by seasonal changes in environmental conditions and human behaviors. Previous studies have shown that associations between temperature and viral infections may be primarily driven by changes in human behavior rather than just meteorological or environmental conditions.^3,5,17,71^ In this coastal, urban setting, the effects of temperature may be less impacted by the immediate effects on pathogen survival and transmissibility but may be more closely aligned with long-term seasonal changes in environmental conditions and human behaviors.

A key strength of this study is the inclusion of enteric pathogen infection data, which was able to show taxa– and pathogen-specific differences in infection patterns. While previous assessments of the PAASIM data showed limited evidence of an association between meteorological conditions and diarrhea [Kann et al., *manuscript in preparation*], here we show evidence of taxa-specific differences in transmission pathways, highlighting the value of enteric pathogen data for assessing these relationships. Though many previous studies have relied on reported diarrhea to study these relationships,^4,8,38,39,41,42,72,73^ the increasing use of enteric pathogen infection data has shown the importance of identifying taxa-level differences in transmission patterns across different contexts.^2,15,16,25,27,74^ Our assessment also shows some evidence of pathogen-specific differences in the effects of meteorological conditions. For example, we saw an association between dry season and HREs (at a 1-week lag period) and a higher prevalence of *Cryptosporidium*. Previous analyses in the PAASIM study area showed that *Cryptosporidium* was present in the water supply [Linden et al., *manuscript in preparation*] and that the intervention was associated with a lower risk of *Cryptosporidium* infections.^66^ The known presence of *Cryptosporidium* in the study area and our finding of a strong seasonal and weather-dependent trend in infections suggests that *Cryptosporidium* may be an important priority for targeted interventions in this study area. Enteric pathogen data, rather than just reported diarrhea, provides important insight into taxa-and pathogen specific transmission patterns and should continue to be collected in similar studies.

This analysis has some limitations. First, our outcome measure was limited in this study by only being an indicator of prevalence rather than incidence of infection. Different pathogens have different levels of persistence in the gut based on a variety of factors (i.e., viral pathogens may persist in the gut for shorter periods than bacterial or protozoan infection).^62^ The assessment of associations with more immediate exposures (e.g. HREs) in particular may be limited because persistent pathogen infections may be shed and detected in stool for long periods. These persistent infections likely are not representative of infections associated with short-term exposures in the lag periods assessed in this study. This may have introduced non-differential misclassification of the outcome and potentially biased results towards the null. We also only had one cross-sectional sample per child and no repeated samples or longitudinal follow-up of individual children, making our analysis reliant on population-level trends in enteric infections rather than within-child variability. Follow up studies could explore trends in enteric infection rates in the study area across different age groups and with a longitudinal follow-up study design. Additionally, data was collected for under a two-year period. While some previous studies have used similar follow up periods,^15,39^ data collected over a longer period may be helpful in future studies to observe long term trends in seasonal infection patterns. We also only had data available for children at 12-months old, which is a limited age range compared to some previous studies that have assessed a range of ages up to 5 years old.^2,15,27,28^ This may also explain some of the differences we saw in seasonal trends of enteric disease compared to other studies. Finally, weather data was collected at a weather station at the Beira airport, located approximately seven kilometers from our study neighborhood. Previous assessments of spatiotemporal differences in rainfall data and study populations may bias results towards the null, though the biasing effect of rain gauge location occurred at distances larger than what we assessed in this study.^75^ Future assessments of weather and enteric disease should consider meteorological exposure estimations that are spatially closer to the study population.

## Conclusions

In this low-income, urban setting we saw distinct seasonal and weather dependent trends in enteric pathogen infections among 12-month-old children. Though observed associations of season and temperature on enteric infections were different than expected based on previous literature, they highlight important long-term changes in environmental conditions and behaviors. Heavy rainfall events represent a more proximal exposure leading to a higher risk of some key enteric infections. Understanding both long-term seasonal variation and the more immediate impacts of specific weather events, such as HREs, is important for developing more resilient intervention strategies across different settings.

## Data Availability

Deidentified data and analysis code can be accessed on our project OSF (Open Science Framework) site upon publication at this link – https://osf.io/f7w5g/

## Acknowledgements

This research was supported by the National Institute of Allergy and Infectious Diseases (NIAID) through grant number R01AI130163. R.S.K. was supported by NIAID through grant number F31AI183829-01 and by the Future Rivers program at the University of Washington as part of an NSF National Research Traineeship award (DGE 1922004). Finally, we would like to thank everyone that contributed to the PAASIM study, including our study participants who gave their time and data for this research.

## The PAASIM Study Authorship Consortium

Thomas Clasen^1^, Konstantinos T. Konstantinidis^4^, Magalhães Mangamela^6^, Sydney Hubbard^1^, Joaquim Domingos Lequechane^5^, Molly K. Miller-Petrie^2^, Lilly A. O’Brien^1^, Courtney P. Victor^1^, Nicolette Angela Zhou^2^, Toheedat Bahara^3^

^1^Emory University, Rollins School of Public Health, Gangarosa Department of Environmental Health, Atlanta, Georgia, USA

^2^University of Washington, Department of Environmental and Occupational Health Sciences, Seattle, WA, USA

^3^University of North Carolina, Gillings School of Public Health, Department of Environmental Sciences and Engineering, Chapel Hilll, NC, USA

^4^Georgia Institute of Technology, School of Civil & Environmental Engineering, Atlanta, GA, USA

^5^Beira Operations Research Center, Beira, Mozambique

^6^AURA, IP, Maputo, Mozambique

## Notes

### Competing Interest Statement

The authors have declared no competing interest.

### Author Declarations

The National Bio-Ethics Committee for Health of Mozambique and the Institutional Review Board of Emory University gave ethical approval for this work

