## Supplemental Materials for "Variability of enteric pathogen infections by season and meteorological conditions in a low-income, urban setting in Mozambique"

**Supplemental Table 1.** Study population characteristics (N=630)

| | n (%) or Mean (SD) |
|---|---|
| Female index child | 302 (47.9%) |
| High poverty based on socio-economic status* | 383 (60.8%) |
| Caregiver completed at least secondary education* | 151 (24.0%) |
| Primary caregiver has fixed employment* | 229 (36.3%) |
| Number of children under 5 living in household* | 1.4 (0.6) |
| Number of people in the household* | 5.6 (2.5) |
| Months living in the household* | 69.7 (80.8) |
| Human feces observed in or near the household* | 6 (1.0%) |
| Animal feces observed in or near the household* | 84 (13.3%) |
| Severely food insecure* | 297 (47.1%) |
| Handwashing station in household or yard at baseline | 129 (20.5%) |
| Basic household sanitation access at baseline | 226 (35.9%) |
| Improved water | 630 (100.0%) |
| Drinking water source on premises | 305 (48.4%) |
| Water insecure (HWISE) at baseline | 86 (13.8%) |

* Data are reported for 12-month visit whereas they are reported at baseline for the main PAASIM analysis

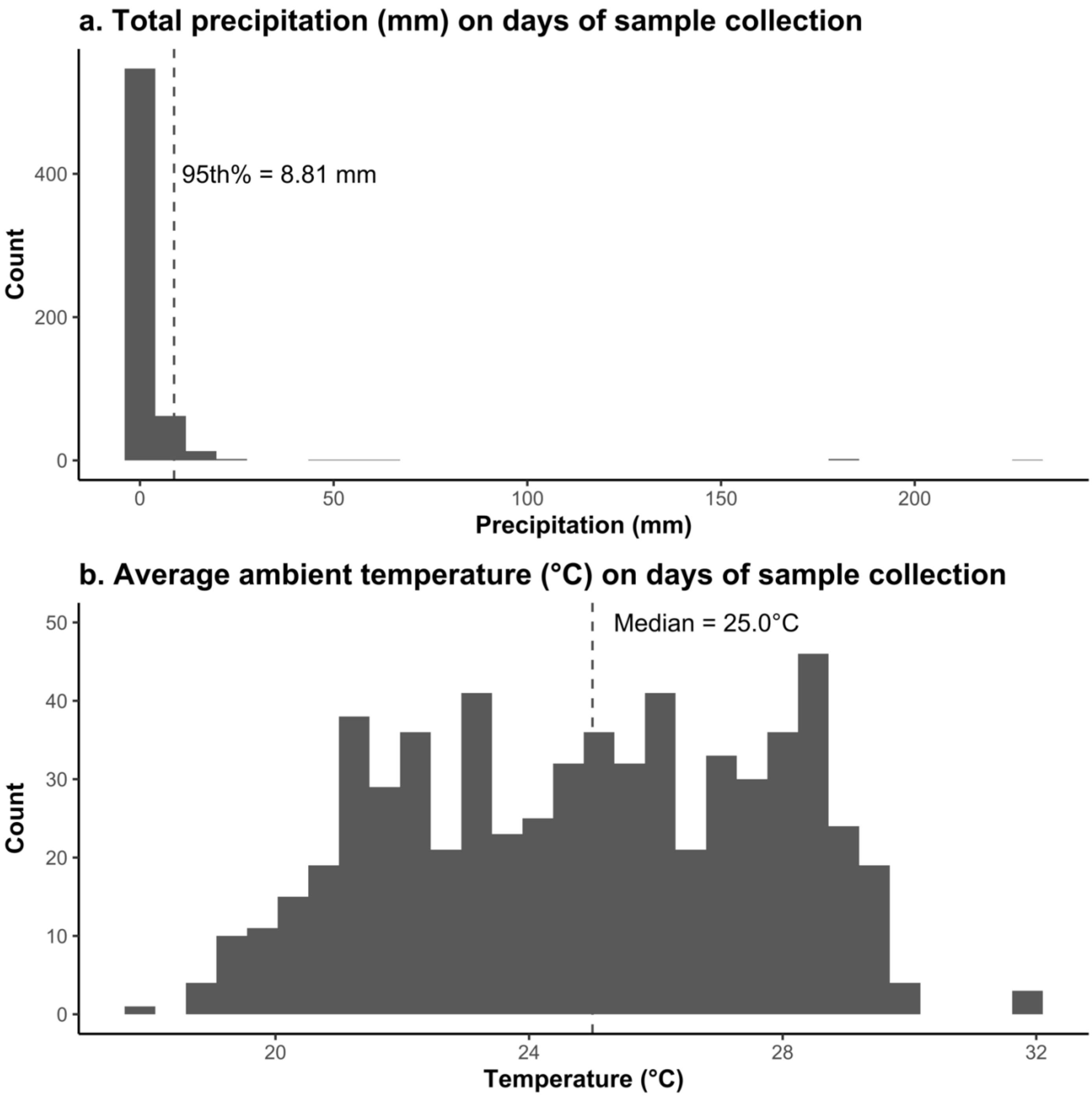


**Supplemental Figure 1.** Distribution of (a) precipitation and (b) temperature on days where stools samples were collected (N=630).

**Supplemental Table 2.** Adjusted associations of Heavy Rainfall Events (HREs) on enteric pathogen infections.

| | **0-1 week before sample** | | **1-2 weeks before sample** | | **2-3 weeks before sample** | |
|---|---|---|---|---|---|---|
| | **aPR or aβ (95% CI)** | ***p-value*** | **aPR or aβ (95% CI)** | ***p-value*** | **aPR or aβ (95% CI)** | ***p-value*** |
| **Combined outcomes** | | | | | | |
| **Any bacteria** | 0.98 (0.88, 1.08) | 0.64 | 1.01 (0.92, 1.10) | 0.85 | 0.99 (0.88, 1.11) | 0.82 |
| **Any protozoa** | 0.93 (0.71, 1.22) | 0.61 | 1.30 (1.06, 1.59) | 0.01 | 0.99 (0.79, 1.24) | 0.93 |
| **Any virus** | 1.24 (0.97, 1.60) | 0.09 | 1.22 (0.95, 1.57) | 0.12 | 0.96 (0.75, 1.22) | 0.73 |
| **Co-infection** | 1.05 (0.92, 1.20) | 0.44 | 1.09 (0.99, 1.21) | 0.09 | 0.99 (0.84, 1.17) | 0.91 |
| **Number of infections** | -0.12 (-0.32, 0.08) | 0.25 | -0.04 (-0.22, 0.14) | 0.63 | -0.01 (-0.24, 0.21) | 0.91 |
| **Bacterial outcomes** | | | | | | |
| **EAEC** | 1.03 (0.85, 1.24) | 0.78 | 0.88 (0.72, 1.08) | 0.23 | 1.09 (0.89, 1.33) | 0.40 |
| **DAEC** | 0.92 (0.82, 1.03) | 0.13 | 1.03 (0.96, 1.12) | 0.42 | 0.94 (0.85, 1.04) | 0.24 |
| **tEPEC** | 0.85 (0.63, 1.14) | 0.27 | 0.94 (0.66, 1.32) | 0.71 | 1.01 (0.63, 1.60) | 0.98 |
| **aEPEC** | 1.06 (0.87, 1.29) | 0.56 | 1.01 (0.79, 1.30) | 0.93 | 0.97 (0.72, 1.32) | 0.86 |
| **ETEC** | 0.83 (0.51, 1.34) | 0.44 | 0.41 (0.23, 0.73) | 0.00 | 0.99 (0.58, 1.68) | 0.96 |
| **Shigella** | 0.90 (0.61, 1.33) | 0.60 | 0.80 (0.56, 1.16) | 0.24 | 1.01 (0.68, 1.50) | 0.95 |
| **Campylobacter** | 0.86 (0.63, 1.18) | 0.35 | 1.23 (0.92, 1.66) | 0.17 | 1.04 (0.73, 1.47) | 0.85 |
| **Viral outcomes** | | | | | | |
| **Norovirus** | 1.62 (1.02, 2.58) | 0.04 | 1.12 (0.68, 1.86) | 0.65 | 0.73 (0.4, 1.33) | 0.30 |
| **Protozoan outcomes** | | | | | | |
| **Cryptosporidium** | 0.70 (0.43, 1.14) | 0.15 | 1.41 (0.98, 2.02) | 0.06 | 1.15 (0.80, 1.65) | 0.46 |
| **Giardia** | 1.12 (0.75, 1.68) | 0.57 | 1.12 (0.84, 1.50) | 0.45 | 0.93 (0.66, 1.31) | 0.67 |

HREs were defined as a day where the total rainfall was above the $95^{th}$ percentile (8.81mm) for the overall study period. All models adjusted for rolling mean temperature during the same period, intervention status, access to a direct household connection to a piped water source, poverty, caregiver education level, caregiver employment status, and basic sanitation access. Models for specific infection only run for enteric infections with prevalence over 10%.

**Supplemental Table 3.** Sensitivity analysis using 80th and 90th percentile HRE cutoffs

| | 0-1 week before sample | | 1-2 weeks before sample | | 2-3 weeks before sample | |
|---|---|---|---|---|---|---|
| | **aPR or aβ (95% CI)** | ***p-value*** | **aPR or aβ (95% CI)** | ***p-value*** | **aPR or aβ (95% CI)** | ***p-value*** |
| **90th Percentile** | | | | | | |
| **Combined outcomes** | | | | | | |
| **Any bacteria** | 0.93 (0.85, 1.03) | 0.15 | 0.93 (0.86, 1.01) | 0.10 | 1.01 (0.90, 1.14) | 0.81 |
| **Any protozoa** | 1.00 (0.79, 1.26) | 0.98 | 1.17 (0.96, 1.43) | 0.11 | 0.99 (0.83, 1.17) | 0.89 |
| **Any virus** | 1.13 (0.91, 1.41) | 0.26 | 1.15 (0.93, 1.41) | 0.19 | 0.94 (0.76, 1.16) | 0.54 |
| **Co-infection** | 0.99 (0.88, 1.11) | 0.83 | 1.01 (0.90, 1.14) | 0.85 | 1.05 (0.89, 1.24) | 0.58 |
| **Number of pathogens** | -0.13 (-0.34, 0.08) | 0.22 | -0.17 (-0.34, 0.00) | 0.08 | 0.02 (-0.20, 0.24) | 0.85 |
| **Bacterial outcomes** | | | | | | |
| **EAEC** | 0.96 (0.82, 1.14) | 0.65 | 0.99 (0.83, 1.17) | 0.89 | 0.97 (0.80, 1.18) | 0.74 |
| **DAEC** | 0.88 (0.79, 0.97) | 0.01 | 0.94 (0.86, 1.02) | 0.15 | 0.94 (0.87, 1.03) | 0.18 |
| **tEPEC** | 0.77 (0.59, 0.99) | 0.04 | 0.89 (0.70, 1.14) | 0.35 | 0.87 (0.60, 1.28) | 0.49 |
| **aEPEC** | 0.97 (0.80, 1.19) | 0.79 | 0.98 (0.79, 1.22) | 0.88 | 1.19 (0.94, 1.51) | 0.14 |
| **ETEC** | 1.08 (0.71, 1.63) | 0.74 | 0.41 (0.26, 0.66) | 0.00 | 0.72 (0.47, 1.10) | 0.12 |
| **Shigella** | 0.95 (0.70, 1.30) | 0.76 | 0.69 (0.49, 0.99) | 0.05 | 1.06 (0.70, 1.61) | 0.77 |
| **Campylobacter** | 0.91 (0.70, 1.18) | 0.46 | 1.18 (0.91, 1.53) | 0.22 | 1.02 (0.76, 1.36) | 0.90 |
| **Viral outcomes** | | | | | | |
| **Norovirus** | 1.44 (0.93, 2.21) | 0.10 | 1.17 (0.74, 1.84) | 0.51 | 0.97 (0.64, 1.46) | 0.87 |
| **Protozoan outcomes** | | | | | | |
| **Cryptosporidium** | 0.79 (0.52, 1.20) | 0.26 | 1.17 (0.80, 1.70) | 0.41 | 1.03 (0.79, 1.34) | 0.84 |
| **Giardia** | 1.10 (0.75, 1.63) | 0.63 | 1.00 (0.77, 1.30) | 0.99 | 1.01 (0.79, 1.29) | 0.95 |
| **80th Percentile** | | | | | | |
| **Combined outcomes** | | | | | | |
| **Any bacteria** | 0.96 (0.85, 1.08) | 0.47 | 0.92 (0.83, 1.03) | 0.15 | 0.96 (0.87, 1.06) | 0.42 |
| **Any protozoa** | 0.77 (0.61, 0.96) | 0.02 | 0.91 (0.72, 1.16) | 0.45 | 0.99 (0.82, 1.20) | 0.93 |
| **Any virus** | 0.97 (0.79, 1.20) | 0.80 | 1.14 (0.90, 1.45) | 0.28 | 0.91 (0.72, 1.14) | 0.39 |
| **Co-infection** | 0.88 (0.75, 1.04) | 0.13 | 0.94 (0.82, 1.08) | 0.38 | 0.96 (0.83, 1.10) | 0.53 |
| **Number of pathogens** | -0.21 (-0.45, 0.04) | 0.10 | -0.15 (-0.36, 0.05) | 0.14 | -0.04 (-0.26, 0.17) | 0.67 |
| **Bacterial outcomes** | | | | | | |
| **EAEC** | 0.95 (0.83, 1.10) | 0.52 | 0.87 (0.75, 1.02) | 0.08 | 0.84 (0.67, 1.05) | 0.12 |
| **DAEC** | 0.93 (0.85, 1.02) | 0.14 | 0.93 (0.84, 1.03) | 0.16 | 0.95 (0.86, 1.05) | 0.31 |

| | | | | | | |
|---|---|---|---|---|---|---|
| **tEPEC** | 0.89 (0.64, 1.22) | 0.46 | 0.85 (0.61, 1.20) | 0.36 | 0.72 (0.49, 1.05) | 0.09 |
| **aEPEC** | 0.84 (0.67, 1.06) | 0.14 | 1.00 (0.79, 1.27) | 1.00 | 1.14 (0.87, 1.48) | 0.35 |
| **ETEC** | 0.92 (0.58, 1.44) | 0.71 | 0.60 (0.34, 1.07) | 0.08 | 0.76 (0.45, 1.26) | 0.28 |
| **Shigella** | 1.05 (0.68, 1.62) | 0.82 | 0.90 (0.62, 1.33) | 0.60 | 0.95 (0.65, 1.37) | 0.77 |
| **Campylobacter** | 1.02 (0.75, 1.38) | 0.93 | 1.08 (0.79, 1.47) | 0.63 | 0.92 (0.68, 1.24) | 0.58 |
| **Viral outcomes** | | | | | | |
| **Norovirus** | 1.20 (0.78, 1.86) | 0.41 | 1.23 (0.76, 2.01) | 0.40 | 1.12 (0.72, 1.73) | 0.62 |
| **Protozoan outcomes** | | | | | | |
| **Cryptosporidium** | 0.65 (0.46, 0.92) | 0.02 | 0.91 (0.62, 1.33) | 0.63 | 1.11 (0.80, 1.56) | 0.54 |
| **Giardia** | 0.83 (0.59, 1.18) | 0.30 | 0.88 (0.65, 1.20) | 0.42 | 1.05 (0.77, 1.44) | 0.74 |

**Supplemental Table 4.** Adjusted associations of Heavy Rainfall Events (HREs) on enteric pathogen infections by enteric pathogen infections including interaction by antecedent conditions.

| | 0-1 week before sample | | 1-2 weeks before sample | | 2-3 weeks before sample | |
|---|---|---|---|---|---|---|
| | **aPR or aβ (95% CI)** | ***p-value*** | **aPR or aβ (95% CI)** | ***p-value*** | **aPR or aβ (95% CI)** | ***p-value*** |
| **Any Bacteria** | | | | | | |
| **Wet** | 0.96 (0.82, 1.11) | 0.57 | 1.06 (0.92, 1.23) | 0.40 | 1.01 (0.87, 1.18) | 0.89 |
| **Medium/Dry** | 1.01 (0.77, 1.34) | 0.92 | 0.98 (0.76, 1.25) | 0.86 | 0.98 (0.75, 1.28) | 0.88 |
| **Interaction** | 1.06 (0.84, 1.34) | 0.63 | 0.92 (0.75, 1.12) | 0.40 | 0.97 (0.78, 1.20) | 0.78 |
| **Any Protozoa** | | | | | | |
| **Wet** | 0.89 (0.59, 1.34) | 0.57 | 1.34 (0.94, 1.93) | 0.11 | 1.09 (0.74, 1.60) | 0.67 |
| **Medium/Dry** | 0.94 (0.49, 1.84) | 0.87 | 1.32 (0.72, 2.41) | 0.37 | 0.85 (0.41, 1.76) | 0.66 |
| **Interaction** | 1.06 (0.63, 1.79) | 0.82 | 0.98 (0.60, 1.59) | 0.94 | 0.78 (0.42, 1.45) | 0.44 |
| **Any Virus** | | | | | | |
| **Wet** | 1.48 (1.12, 1.94) | 0.01 | 1.24 (0.95, 1.63) | 0.12 | 1.30 (0.97, 1.74) | 0.08 |
| **Medium/Dry** | 1.03 (0.59, 1.80) | 0.92 | 1.25 (0.79, 1.98) | 0.34 | 0.64 (0.35, 1.16) | 0.14 |
| **Interaction** | 0.70 (0.43, 1.13) | 0.15 | 1.01 (0.69, 1.46) | 0.97 | 0.49 (0.29, 0.83) | 0.01 |
| **Co-infection** | | | | | | |
| **Wet** | 1.02 (0.85, 1.23) | 0.82 | 1.14 (0.95, 1.37) | 0.16 | 1.08 (0.88, 1.32) | 0.48 |
| **Medium/Dry** | 1.08 (0.76, 1.54) | 0.66 | 1.04 (0.75, 1.43) | 0.82 | 0.89 (0.62, 1.26) | 0.50 |
| **Interaction** | 1.06 (0.78, 1.43) | 0.71 | 0.91 (0.70, 1.19) | 0.50 | 0.82 (0.62, 1.10) | 0.19 |
| **Number of Pathogens** | | | | | | |
| **Wet** | -0.17 (-0.45, 0.11) | 0.24 | 0.01 (-0.25, 0.27) | 0.94 | 0.04 (-0.25, 0.32) | 0.80 |
| **Medium/Dry** | -0.08 (-0.56, 0.41) | 0.76 | -0.14 (-0.54, 0.27) | 0.50 | -0.08 (-0.57, 0.41) | 0.75 |
| **Interaction** | 0.09 (-0.30, 0.49) | 0.65 | -0.15 (-0.46, 0.17) | 0.36 | -0.12 (-0.51, 0.28) | 0.56 |
| **EAEC** | | | | | | |
| **Wet** | 0.91 (0.70, 1.19) | 0.51 | 0.95 (0.72, 1.25) | 0.71 | 1.22 (0.93, 1.61) | 0.14 |
| **Medium/Dry** | 1.22 (0.72, 2.06) | 0.45 | 0.89 (0.56, 1.39) | 0.60 | 1.05 (0.65, 1.69) | 0.84 |
| **Interaction** | 1.34 (0.85, 2.10) | 0.21 | 0.93 (0.65, 1.33) | 0.70 | 0.86 (0.58, 1.27) | 0.45 |
| **DAEC** | | | | | | |
| **Wet** | 0.92 (0.81, 1.06) | 0.24 | 0.99 (0.86, 1.13) | 0.85 | 1.06 (0.93, 1.22) | 0.38 |
| **Medium/Dry** | 0.92 (0.72, 1.17) | 0.50 | 1.10 (0.87, 1.37) | 0.43 | 0.81 (0.63, 1.02) | 0.08 |
| **Interaction** | 1.00 (0.81, 1.22) | 0.97 | 1.11 (0.93, 1.33) | 0.26 | 0.76 (0.62, 0.92) | 0.01 |
| **aEPEC** | | | | | | |
| **Wet** | 1.01 (0.73, 1.40) | 0.96 | 1.20 (0.88, 1.64) | 0.26 | 1.08 (0.70, 1.65) | 0.74 |
| **Medium/Dry** | 1.17 (0.64, 2.14) | 0.62 | 0.87 (0.51, 1.5) | 0.62 | 0.89 (0.46, 1.73) | 0.74 |
| **Interaction** | 1.16 (0.70, 1.92) | 0.57 | 0.73 (0.47, 1.14) | 0.16 | 0.83 (0.50, 1.37) | 0.47 |

| | | | | | | |
|---|---|---|---|---|---|---|
| **tEPEC** | | | | | | |
| **Wet** | 0.89 (0.59, 1.32) | 0.55 | 1.00 (0.61, 1.64) | 1.00 | 0.97 (0.55, 1.73) | 0.92 |
| **Medium/Dry** | 0.79 (0.35, 1.82) | 0.59 | 0.84 (0.34, 2.06) | 0.70 | 1.09 (0.45, 2.67) | 0.85 |
| **Interaction** | 0.90 (0.43, 1.86) | 0.77 | 0.84 (0.40, 1.78) | 0.65 | 1.12 (0.57, 2.23) | 0.74 |
| **ETEC** | | | | | | |
| **Wet** | 0.79 (0.31, 2.05) | 0.63 | 0.77 (0.33, 1.81) | 0.55 | 1.53 (0.63, 3.70) | 0.35 |
| **Medium/Dry** | 1.06 (0.21, 5.27) | 0.95 | 0.21 (0.03, 1.35) | 0.10 | 0.93 (0.22, 3.98) | 0.92 |
| **Interaction** | 1.33 (0.36, 4.88) | 0.67 | 0.27 (0.05, 1.42) | 0.12 | 0.61 (0.19, 1.93) | 0.40 |
| **Shigella** | | | | | | |
| **Wet** | 0.90 (0.52, 1.56) | 0.71 | 0.65 (0.4, 1.06) | 0.08 | 1.06 (0.63, 1.79) | 0.82 |
| **Medium/Dry** | 0.86 (0.33, 2.25) | 0.76 | 0.97 (0.37, 2.54) | 0.95 | 0.93 (0.38, 2.28) | 0.87 |
| **Interaction** | 0.95 (0.43, 2.10) | 0.90 | 1.5 (0.65, 3.43) | 0.34 | 0.88 (0.42, 1.81) | 0.72 |
| **Campylobacter** | | | | | | |
| **Wet** | 0.82 (0.51, 1.32) | 0.42 | 1.32 (0.87, 2.01) | 0.19 | 1.08 (0.69, 1.70) | 0.73 |
| **Medium/Dry** | 0.85 (0.36, 1.99) | 0.70 | 1.04 (0.53, 2.03) | 0.90 | 0.83 (0.39, 1.76) | 0.63 |
| **Interaction** | 1.03 (0.50, 2.09) | 0.94 | 0.79 (0.47, 1.33) | 0.37 | 0.77 (0.42, 1.4) | 0.38 |
| **Norovirus** | | | | | | |
| **Wet** | 2.26 (1.22, 4.21) | 0.01 | 1.08 (0.63, 1.85) | 0.79 | 0.67 (0.26, 1.74) | 0.41 |
| **Medium/Dry** | 1.49 (0.54, 4.08) | 0.44 | 1.46 (0.54, 3.92) | 0.45 | 0.91 (0.19, 4.41) | 0.90 |
| **Interaction** | 0.66 (0.30, 1.46) | 0.30 | 1.36 (0.59, 3.11) | 0.47 | 1.35 (0.38, 4.78) | 0.64 |
| **Cryptosporidium** | | | | | | |
| **Wet** | 0.64 (0.33, 1.23) | 0.18 | 1.60 (0.86, 2.99) | 0.14 | 1.34 (0.76, 2.34) | 0.31 |
| **Medium/Dry** | 0.74 (0.28, 1.93) | 0.53 | 1.25 (0.45, 3.50) | 0.67 | 0.91 (0.32, 2.57) | 0.86 |
| **Interaction** | 1.16 (0.57, 2.32) | 0.69 | 0.78 (0.35, 1.77) | 0.56 | 0.68 (0.28, 1.63) | 0.39 |
| **Giardia** | | | | | | |
| **Wet** | 1.13 (0.62, 2.08) | 0.69 | 1.10 (0.68, 1.77) | 0.71 | 0.87 (0.51, 1.47) | 0.60 |
| **Medium/Dry** | 1.11 (0.38, 3.19) | 0.85 | 1.11 (0.44, 2.80) | 0.83 | 0.97 (0.40, 2.40) | 0.95 |
| **Interaction** | 0.98 (0.41, 2.33) | 0.96 | 1.01 (0.46, 2.23) | 0.98 | 1.12 (0.54, 2.34) | 0.76 |

Antecedent conditions were calculated using the sum of total rainfall over the 8 weeks prior to sample collection, where wet conditions are those exceeding 67$^{th}$ percentile for total rainfall over the 8-week period compared to the study period and medium/dry conditions are those below the 67$^{th}$ percentile for total rainfall. HREs were defined as a day where the total rainfall was above the 95$^{th}$ percentile (8.81mm) for the overall study period. All models adjusted for rolling mean temperature during the same period, intervention status, access to a direct household connection to a piped water source, poverty, caregiver education level, caregiver employment status, and basic sanitation access.

**Supplemental Table 5.** Adjusted associations of above median temperatures on enteric pathogen infections.

| | **0-1 week before sample** | | **1-2 weeks before sample** | | **2-3 weeks before sample** | |
|---|---|---|---|---|---|---|
| | **aPR or aβ (95% CI)** | ***p-value*** | **aPR or aβ (95% CI)** | ***p-value*** | **aPR or aβ (95% CI)** | ***p-value*** |
| **Combined outcomes** | | | | | | |
| **Any bacteria** | 1.00 (0.91, 1.10) | 0.99 | 0.98 (0.89, 1.07) | 0.61 | 0.99 (0.90, 1.10) | 0.91 |
| **Any protozoa** | 0.62 (0.47, 0.80) | <0.01 | 0.65 (0.49, 0.86) | <0.01 | 0.62 (0.48, 0.79) | 0.00 |
| **Any virus** | 0.96 (0.76, 1.23) | 0.77 | 0.94 (0.74, 1.19) | 0.62 | 1.05 (0.82, 1.34) | 0.70 |
| **Co-infection** | 0.91 (0.80, 1.04) | 0.17 | 0.86 (0.76, 0.97) | 0.02 | 0.92 (0.81, 1.03) | 0.16 |
| **Number of infections** | -0.11 (-0.33, 0.11) | 0.33 | -0.13 (-0.33, 0.07) | 0.19 | -0.14 (-0.36, 0.08) | 0.20 |
| **Bacterial outcomes** | | | | | | |
| **EAEC** | 1.08 (0.92, 1.26) | 0.34 | 1.08 (0.91, 1.29) | 0.36 | 1.16 (0.96, 1.40) | 0.13 |
| **DAEC** | 0.94 (0.85, 1.03) | 0.19 | 0.89 (0.80, 0.98) | 0.01 | 0.91 (0.82, 1.01) | 0.07 |
| **tEPEC** | 1.15 (0.82, 1.62) | 0.41 | 1.10 (0.77, 1.58) | 0.59 | 1.07 (0.76, 1.50) | 0.72 |
| **aEPEC** | 0.93 (0.76, 1.14) | 0.49 | 0.87 (0.71, 1.07) | 0.19 | 0.93 (0.76, 1.13) | 0.46 |
| **ETEC** | 1.49 (0.92, 2.42) | 0.11 | 1.45 (0.86, 2.43) | 0.16 | 1.22 (0.72, 2.07) | 0.46 |
| **Shigella** | 1.23 (0.80, 1.90) | 0.35 | 1.24 (0.79, 1.94) | 0.35 | 1.21 (0.79, 1.87) | 0.38 |
| **Campylobacter** | 1.01 (0.74, 1.38) | 0.96 | 0.91 (0.70, 1.18) | 0.46 | 1.00 (0.77, 1.30) | 1.00 |
| **Viral outcomes** | | | | | | |
| **Norovirus** | 1.44 (0.89, 2.33) | 0.13 | 1.41 (0.89, 2.21) | 0.14 | 1.65 (1.08, 2.54) | 0.02 |
| **Protozoan infections** | | | | | | |
| **Cryptosporidium** | 0.43 (0.27, 0.68) | <0.01 | 0.40 (0.27, 0.59) | <0.01 | 0.38 (0.27, 0.54) | <0.01 |
| **Giardia** | 0.77 (0.56, 1.07) | 0.12 | 0.93 (0.64, 1.37) | 0.72 | 0.87 (0.62, 1.20) | 0.39 |

Above median temperatures were defined as rolling average weekly temperature above the 50th percentile (25.0ºC) for the full study period. All models adjusted for rolling mean precipitation during the same period, intervention status, access to a direct household connection to a piped water source, poverty, caregiver education level, caregiver employment status, and basic sanitation access. Models only run for enteric infections with prevalence over 10%.

**Supplemental Table 6.** Adjusted associations of below 33rd tertile temperatures on enteric pathogen infections.

| | **0-1 week before sample** | | **1-2 weeks before sample** | | **2-3 weeks before sample** | |
|---|---|---|---|---|---|---|
| | **aPR or aβ (95% CI)** | ***p-value*** | **aPR or aβ (95% CI)** | ***p-value*** | **aPR or aβ (95% CI)** | ***p-value*** |
| **Combined outcomes** | | | | | | |
| **Any bacteria** | 1.02 (0.94, 1.12) | 0.59 | 0.98 (0.89, 1.07) | 0.65 | 0.99 (0.90, 1.10) | 0.91 |
| **Any protozoa** | 1.47 (1.16, 1.85) | 0.00 | 1.42 (1.12, 1.80) | 0.00 | 1.43 (1.09, 1.87) | 0.01 |
| **Any virus** | 0.91 (0.74, 1.12) | 0.35 | 0.83 (0.65, 1.06) | 0.13 | 0.93 (0.73, 1.18) | 0.54 |
| **Co-infection** | 1.16 (1.02, 1.31) | 0.02 | 1.09 (0.96, 1.23) | 0.18 | 1.13 (0.98, 1.30) | 0.09 |
| **Number of infections** | 0.15 (-0.04, 0.35) | 0.12 | 0.11 (-0.09, 0.32) | 0.27 | 0.12 (-0.11, 0.35) | 0.30 |
| **Bacterial outcomes** | | | | | | |
| **EAEC** | 0.92 (0.75, 1.12) | 0.41 | 0.84 (0.70, 1.00) | 0.05 | 0.94 (0.78, 1.13) | 0.47 |
| **DAEC** | 1.04 (0.94, 1.14) | 0.48 | 1.09 (1.01, 1.19) | 0.03 | 1.09 (0.99, 1.18) | 0.07 |
| **tEPEC** | 0.79 (0.57, 1.09) | 0.15 | 0.82 (0.59, 1.16) | 0.27 | 0.75 (0.53, 1.04) | 0.08 |
| **aEPEC** | 1.02 (0.85, 1.23) | 0.83 | 1.13 (0.91, 1.41) | 0.26 | 1.14 (0.92, 1.43) | 0.24 |
| **ETEC** | 0.87 (0.52, 1.45) | 0.59 | 0.73 (0.43, 1.24) | 0.24 | 0.76 (0.43, 1.33) | 0.33 |
| **Shigella** | 1.27 (0.83, 1.94) | 0.26 | 0.98 (0.62, 1.54) | 0.92 | 0.93 (0.57, 1.52) | 0.76 |
| **Campylobacter** | 1.09 (0.84, 1.41) | 0.52 | 1.03 (0.81, 1.31) | 0.80 | 0.94 (0.75, 1.19) | 0.61 |
| **Viral outcomes** | | | | | | |
| **Norovirus** | 0.47 (0.27, 0.83) | 0.01 | 0.51 (0.28, 0.93) | 0.03 | 0.64 (0.37, 1.11) | 0.11 |
| **Protozoan infections** | | | | | | |
| **Cryptosporidium** | 1.99 (1.41, 2.82) | 0.00 | 1.91 (1.35, 2.71) | 0.00 | 1.90 (1.34, 2.70) | 0.00 |
| **Giardia** | 1.19 (0.87, 1.62) | 0.28 | 1.11 (0.77, 1.60) | 0.58 | 1.13 (0.80, 1.60) | 0.48 |

Below 33rd percentile temperatures were defined as rolling average weekly temperature below the 33rd percentile (23.3ºC) for the full study period. All models adjusted for rolling mean precipitation during the same period, intervention status, access to a direct household connection to a piped water source, poverty, caregiver education level, caregiver employment status, and basic sanitation access.

**Supplemental Table 7.** Adjusted associations of above 66th tertile temperatures on enteric pathogen infections.

| | **0-1 week before sample** | | **1-2 weeks before sample** | | **2-3 weeks before sample** | |
|---|---|---|---|---|---|---|
| | **aPR or aβ (95% CI)** | ***p-value*** | **aPR or aβ (95% CI)** | ***p-value*** | **aPR or aβ (95% CI)** | ***p-value*** |
| **Combined outcomes** | | | | | | |
| **Any bacteria** | 1.01 (0.92, 1.11) | 0.89 | 0.97 (0.88, 1.07) | 0.56 | 0.99 (0.90, 1.09) | 0.81 |
| **Any protozoa** | 0.55 (0.43, 0.70) | 0.00 | 0.58 (0.44, 0.76) | 0.00 | 0.70 (0.52, 0.93) | 0.01 |
| **Any virus** | 0.86 (0.66, 1.11) | 0.25 | 1.02 (0.79, 1.31) | 0.91 | 0.91 (0.69, 1.22) | 0.53 |
| **Co-infection** | 0.83 (0.73, 0.95) | 0.00 | 0.88 (0.77, 1.02) | 0.09 | 0.89 (0.78, 1.01) | 0.07 |
| **Number of infections** | -0.15 (-0.32, 0.03) | 0.10 | -0.20 (-0.38, -0.02) | 0.03 | -0.15 (-0.32, 0.01) | 0.07 |
| **Bacterial outcomes** | | | | | | |
| **EAEC** | 1.08 (0.93, 1.26) | 0.32 | 0.98 (0.82, 1.16) | 0.78 | 0.92 (0.77, 1.08) | 0.30 |
| **DAEC** | 0.85 (0.76, 0.95) | 0.00 | 0.84 (0.76, 0.94) | 0.00 | 0.88 (0.79, 0.98) | 0.02 |
| **tEPEC** | 1.03 (0.73, 1.46) | 0.85 | 1.06 (0.76, 1.47) | 0.74 | 1.11 (0.78, 1.58) | 0.56 |
| **aEPEC** | 1.03 (0.83, 1.29) | 0.79 | 0.89 (0.74, 1.08) | 0.24 | 0.87 (0.72, 1.05) | 0.14 |
| **ETEC** | 1.36 (0.80, 2.31) | 0.26 | 1.08 (0.64, 1.82) | 0.79 | 1.08 (0.66, 1.77) | 0.77 |
| **Shigella** | 1.10 (0.72, 1.66) | 0.66 | 0.99 (0.66, 1.49) | 0.96 | 1.14 (0.80, 1.62) | 0.47 |
| **Campylobacter** | 1.02 (0.73, 1.44) | 0.89 | 0.99 (0.74, 1.31) | 0.93 | 0.95 (0.70, 1.29) | 0.73 |
| **Viral outcomes** | | | | | | |
| **Norovirus** | 1.63 (1.09, 2.45) | 0.02 | 1.68 (1.06, 2.67) | 0.03 | 1.32 (0.91, 1.90) | 0.14 |
| **Protozoan infections** | | | | | | |
| **Cryptosporidium** | 0.44 (0.28, 0.69) | 0.00 | 0.31 (0.20, 0.50) | 0.00 | 0.40 (0.25, 0.63) | 0.00 |
| **Giardia** | 0.64 (0.45, 0.92) | 0.01 | 0.82 (0.59, 1.15) | 0.26 | 0.95 (0.69, 1.32) | 0.78 |

Above 66th percentile temperatures were defined as rolling average weekly temperature above the 66th percentile (26.3ºC) for the full study period. All models adjusted for rolling mean precipitation during the same period, intervention status, access to a direct household connection to a piped water source, poverty, caregiver education level, caregiver employment status, and basic sanitation access.
